# Exploring evidence of SGLT2 inhibitors use in Southeast Asia: a systematic review

**DOI:** 10.1101/2025.07.02.25330722

**Authors:** Wei Jin Wong, Ying Zhang, Christopher Harrison, Kit Mun Tan, Mark Woodward, Tu Nguyen

## Abstract

**Background:** The benefits of sodium glucose cotransporter 2 (SGLT2) inhibitors are increasingly recognized, not only in the management of diabetes but also in cardiovascular and renal conditions. However, evidence regarding SGLT2 inhibitor use in Southeast Asian populations remains limited and has yet to be systematically reviewed.

**Methods:** A systematic literature search was conducted in Ovid Medline, Cochrane Central Register of Controlled Trials and Embase from database inception until 21 October 2024. Studies were included if they were conducted in adults living in Southeast Asian countries and contained information on the use of SGLT2 inhibitors.

**Results:** A total of 23 studies were included. These studies were conducted in only 5 out of 11 countries in the region (Malaysia, Philippines, Singapore, Thailand and Vietnam). Among these 23 studies, 12 were conducted in patients with type 2 diabetes, 7 in patients with heart failure, and 4 in patients with chronic kidney disease. Thirteen studies focused on economic evaluations. Limited evidence from these 5 countries suggests that SGLT2 inhibitors appear to be safe, effective, and are likely to be cost-effective and cost-saving.

**Conclusions:** Published evidence in Southeast Asian countries showed that SGLT2 inhibitor treatment may offer promising benefits for patients with type 2 diabetes, heart failure and chronic kidney disease. Further research is needed to understand the availability and affordability, as well as the safety, of SGLT2 inhibitors, taking into consideration the effect of ageing and frailty in this region. In addition, key region-specific factors, such as genetic variations, healthcare infrastructure, and cultural considerations remain to be addressed.

## Introduction

Sodium glucose cotransporter 2 (SGLT2) inhibitors have become an important, effective choice of therapy for the treatment and management of diabetes.^1,2^ These medications work by blocking the reabsorption of glucose and sodium in the renal proximal convoluted tubule, which promotes their excretion and helps manage blood glucose levels effectively.^1^ SGLT2 inhibitors were first introduced to clinical practice about a decade ago, beginning with canagliflozin, which received approval from the U.S. Food and Drug Administration in 2013 for the treatment of type 2 diabetes. In 2014, dapagliflozin and empagliflozin were approved, followed by sotagliflozin and bexagliflozin in 2023.^1,2^ Beyond its role in glycemic management, SGLT2 inhibitors have also demonstrated significant cardiovascular and renal benefits.^3–5^ These benefits include a reduction in major adverse cardiovascular events (MACE), hospitalization for heart failure (HF) and delay in progression of chronic kidney disease (CKD) among patients with type 2 diabetes.^6^ In 2019, the Dapagliflozin in Patients with Heart Failure and Reduced Ejection Fraction (DAPA-HF) trial showed that the risk of worsening HF or death from cardiovascular causes was lower among participants who received dapagliflozin treatment compared to placebo.^7^ Published in 2022, the Dapagliflozin in Heart Failure with Mildly Reduced or Preserved Ejection Fraction (DELIVER) trial reported that dapagliflozin reduced the risk of worsening HF or cardiovascular death in patients with HF and a mildly reduced or preserved ejection fraction.^8^ The DELIVER Investigators noted the necessity for further research in other populations, such as ethnically diverse groups.^8^ Similar findings in patients with HF were also reported with another SGLT2 inhibitor, empagliflozin, in the EMPEROR-Preserved trial.^9^ Additionally, SGLT2 inhibitors have also been associated with potential improvements of hepatic steatosis and/or fibrosis in patients with non-alcoholic fatty liver disease.^10^ Along with its glycemic benefits, these additional therapeutic effects make SGLT2 inhibitors an important treatment of choice for diabetes and its associated comorbidities.

While the clinical efficacy and safety of SGLT2 inhibitors have been well-established in Western populations, their use in Southeast Asia warrants further investigation due to the region’s unique demographic, genetic, environmental, and healthcare characteristics. The Southeast Asia region is experiencing an increasing burden of type 2 diabetes, driven by rapid urbanization, changes in dietary habits, and decreasing physical activity.^11^ With Asia being recognized as the epicentre of the global type 2 diabetes mellitus epidemic, the prevalence of diabetes is particularly evident in Southeast Asia.^12^ For example, Indonesia was ranked fifth globally in terms of number of people living with diabetes, with an estimated 19.5 million people in 2021.^13^ Type 2 diabetes was also found to have a greater impact as a comorbidity in patients in the Southeast Asia when compared to those in Western countries. For example, Banks et al. reported that diabetes was 3 times more prevalent in Singaporean patients with HF compared to their Swedish counterparts, and more strongly associated with poor outcomes.^14^ Southeast Asia also has a high burden of CKD^15^ and one of the highest rates of end-stage renal disease (ESRD) in the world.^12^ Countries in the Southeast Asia region are also experiencing an ongoing epidemiological transition with a growing population of older people.^16^ In older people, the presence of geriatric conditions, such as frailty, multimorbidity, malnutrition, polypharmacy, and disability, can significantly complicate the management of diabetes.^17,18^ Concurrently, the region also faces significant healthcare disparities, including varying access to modern medications and healthcare services, especially in rural and lower-income settings.^16^

As identified by O’Hara et al.^19^, further exploration is needed regarding the access to, and use of, SGLT2 inhibitors in low- to middle-income countries and underserved populations. Data examining the use of SGLT2 inhibitors are limited in Southeast Asia. Lifestyle, cultural, social factors and biological traits may influence the uptake and effect of SGLT2 inhibitors. Thus, there is a need for population-specific evaluations in the use of medicines. A recent expert panel statement from Asia-Pacific countries^20^ highlighted the low utilization of SGLT2 inhibitors in many countries, despite the growing evidence of their usefulness, particularly in resource-limited settings. Therefore, this paper seeks to review the existing data on the use of SGLT2 inhibitors in Southeast Asian countries in order to provide an update on how SGLT2 inhibitors are currently used in the region.

## Methods

A systematic electronic literature search was conducted in the following databases from inception until 21 October 2024: (1) Ovid Medline, (2) Cochrane Central register of controlled trials, and (3) Embase. This review followed the PRISMA guidelines.^21^

The term used for search was ‘sodium-glucose transporter 2 inhibitors’, which was then combined with the names of the Southeast Asian countries. For this review, Southeast Asian countries that were included follows the regional grouping in the Association of Southeast Asian Nations (ASEAN). The 11 included countries were (in alphabetical order) Brunei, Cambodia, Indonesia, Laos, Malaysia, Myanmar, Philippines, Singapore, Thailand, Timor Leste and Vietnam.

Details of the search strategy are as below:

#1 (Sodium-Glucose Transporter 2 inhibitors) OR (Sodium glucose co-transporter type 2 inhibitor) OR (Sodium glucose cotransporter 2 inhibitor) OR (Sodium dependent glucose cotransporter 2 inhibitor) OR (SGLT-2 inhibitors) OR (Gliflozins) OR (Gliflozin) OR (Canagliflozin) OR (Empagliflozin) OR (Ertugliflozin) OR (Sotagliflozin) OR (Dapagliflozin) OR (Ipragliflozin) OR (Lusegliflozin) OR (Tofogliflozin)
#2 (Brunei) OR (Burma) OR (Burmese) OR (Cambodia) OR (Cambodian) OR (Indonesia) OR (Indonesian) OR (Laos) OR (Malaysia) OR (Malaysian) OR (Myanmar) OR (Philippines) OR (Filipino) OR (Singapore) OR (Singaporean) OR (Southeast Asia) OR (Thailand) OR (Thai) OR (Timor Leste) OR (Vietnam) OR (Vietnamese)
#3 Search: #1 AND #2 Studies were included if (1) they were conducted in countries in Southeast Asia and (2) explored the use of SGLT2 inhibitors. The exclusion criteria were animal studies, abstract only, case reports, and systematic reviews. Multi-country studies that included Southeast Asian countries as part of their study sites were also excluded if they did not provide results for a specific country in Southeast Asia.

Results from the search were managed using Covidence®. Study titles and abstracts were screened independently by two members of the research team, based on the inclusion and exclusion criteria. The full texts of qualified publications were read and selected for the final decision to include after discussion among these two reviewers. Any disagreement was solved by discussion with the third member of the team.

We extracted information on the study design, country/countries where participants were recruited, the type of SGLT2 inhibitors, study populations (sample size, age, the main disease of interest), study aims and findings. For studies conducted in older adults (aged 60 years or above), we also searched for data related to frailty, an important geriatric syndrome that can affect the safety and efficacy of SGLT2 inhibitors.^22^ The results of this review were summarized narratively.

## Results

### Description of the included studies

A total of 23 eligible studies were included in this review, following the screening of 288 publication records (Figure 1). There were 8 from Thailand^23–30^, 7 from Malaysia^31–37^, 6 from Singapore^38–43^, 1 from Philippines^44^ and 1 study^45^ that was conducted in three Southeast Asian countries (Malaysia, Thailand and Vietnam). Among the included studies, 12 studies were conducted in patients with type 2 diabetes^23–27,31–33,38–41^, 7 studies in patients with heart failure^28,29,34–36,42,44^, and 4 studies in patients with chronic kidney disease^30,37,43,45^. The types of SGLT2 inhibitors that were used in these studies were canagliflozin, dapagliflozin and empagliflozin (Table 1). In terms of study designs, 9 were observational studies^25–27,32,38–41,43^, 1 was a randomized controlled trial^33^ and 13 were economic evaluation studies^23,24,28–31,34–37,42,44,45^. The sample sizes of these observational studies ranged from 57^40^ to 67,556^38^ (with 3 studies^32,40,41^ having less than 100 participants). In terms of age, 5 studies^23,28–30,36^ were conducted in older adults (aged 60 years or above), while the remaining studies were in adult populations. Among the 5 studies conducted in older adults, there was no data related to frailty.

**Figure 1.**
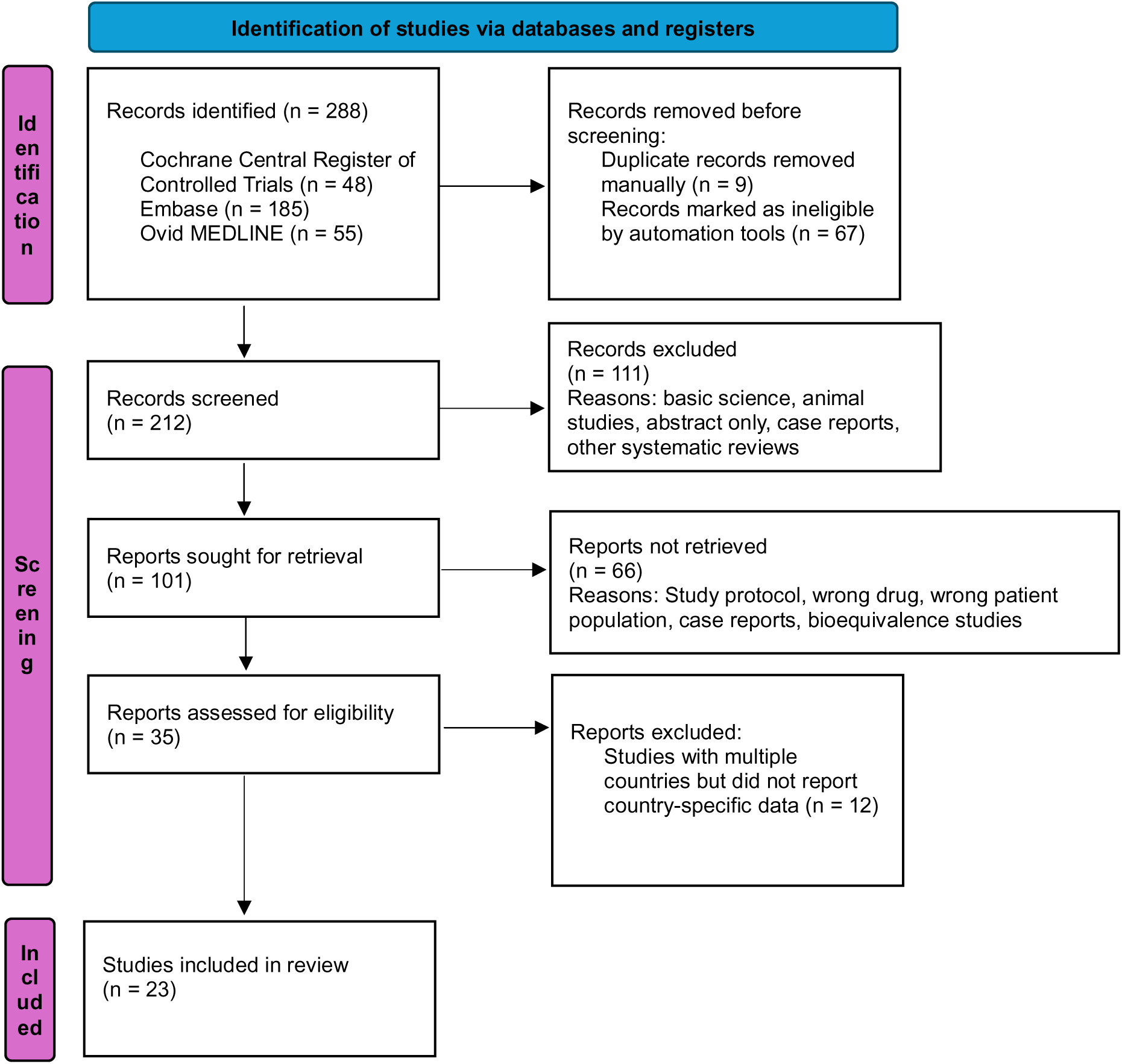
PRISMA flowchart

**Table 1.** Summary of the 23 studies included in this review

| No | Author (Last surname), Year, Title | Study designs | Southeast Asian Countries involved | Types of SGLT2 inhibitors involved | Study Population (sample size and mean/median age) | Aim & Findings |
| --- | --- | --- | --- | --- | --- | --- |
| Type 2 diabetes (12 studies) |  |  |  |  |  |  |
| 1 | Kongmalai, 2024<sup>23</sup><br>Cost-Utility and Budget Impact Analysis of Adding SGLT-2 Inhibitors to Standard Treatment in Type 2 Diabetes Patients with Heart Failure: Utilizing National Database Insights from Thailand | Cost-utility and Budget Impact analysis<br>Societal perspective | Thailand | Canagliflozin, Dapagliflozin, Empagliflozin, | Adult T2DM patients who had previously been diagnosed with HF and exhibited a range of baseline New York Heart Association (NYHA) and left ventricular ejection fraction (LVEF) values.<br>Patients entered the model at 60 years old. | **Aim:**<br>To evaluate the cost-utility of adding SGLT2i to the standard treatment for T2DM-HF patients in Thailand<br>**Findings:**<br>Canagliflozin produced the greatest gain in QALYs, with an increase of 1.21 years, followed by dapagliflozin (0.54 years) and empagliflozin (0.06 years). On average, SGLT2 inhibitors resulted in a 0.41-year gain in QALYs. Canagliflozin also had the highest additional treatment cost at US\$5,600, compared to dapagliflozin at US\$3,547 and empagliflozin at US\$2,694. The ICERs were US\$4,632 for canagliflozin, US\$6,430 for dapagliflozin, US\$48,952 for empagliflozin, and US\$8,480 for SGLT2i overall, per QALY gained. These values exceed Thailand's willingness-to-pay threshold of US\$4,564 per QALY, indicating that SGLT2i treatments are not cost-effective in this setting. Threshold analysis suggests that to meet cost-effectiveness criteria, the prices of canagliflozin, dapagliflozin, empagliflozin, and SGLT2i overall would need to be reduced by 2.3%, 38.2%, 90.2%, and 55.6%, respectively. A budget impact analysis projected a five-year cost of US\$15.6 million for treating type 2 diabetes patients with heart failure. Overall, adding canagliflozin, dapagliflozin, empagliflozin, or any SGLT2i to standard care was likely to not be considered cost-effective for these patients in the Thai healthcare context. |
| 2 | Deerochanawong, 2021<sup>24</sup><br>Cost–Utility Analysis of Dapagliflozin as an Add-On to Standard Treatment for Patients with Type 2 Diabetes and High Risk of Cardiovascular Disease in Thailand | Cost-utility analysis<br>Societal perspective (direct medical and non-medical costs). Indirect costs was not taken into consideration | Thailand | Dapagliflozin | Aged ≥40 years, T2DM, HbA1c in range of 6.5%-12%, CrCl≥60mL/min, have multiple risk factors or established ASCVD | Aim:<br>To analyze the cost–utility of adding dapagliflozin to the standard treatment for treating patients with type 2 diabetes and high cardiovascular risk in a Thai context<br>Findings:<br>The addition of dapagliflozin increased the treatment cost from USD 8,707 to USD 14,455, while improving QALYs from 9.28 to 9.58. This resulted in an ICER of USD 18,988 per QALY. Compared to standard care, patients receiving dapagliflozin experienced greater clinical benefits, including fewer hospitalizations for heart failure and reduced incidence of macroalbuminuria. However, it was not considered cost-effective when assessed against the local cost-effectiveness threshold of 160,000 THB (approximately USD 5,310) per QALY. |
| 3 | Sim, 2023<sup>31</sup> | Cost-effectiveness | Malaysia | SGLT2 inhibitors | Adults with T2DM | Aim:<br>To evaluate the cost-effectiveness of various |
|  | Cost-effectiveness of glucose-lowering therapies as add-on to standard care for people with type 2 diabetes in Malaysia | study<br>Healthcare payer's perspective<br>Model was developed for 4 treatments: Standard care, DPP4i, SGLT2i and GLP1RA |  |  | Patients entered the model at 53 years old. Mean age could not be obtained from the paper | glucose-lowering therapies as add-on to standard care for people with T2DM.<br>Findings:<br>The cost of treating an individual with T2DM ranged from RM 12,494 to RM 41,250, with QALY gains varying between 6.155 and 6.731, depending on the treatment option. Using a willingness-to-pay threshold of RM 29,080 per QALY, SGLT2i emerged as the most cost-effective glucose-lowering therapy when used as an add-on to standard care over a patient's lifetime. This approach yielded a net monetary benefit of RM 176,173 and an ICER of RM 12,279 per QALY gained. Compared to standard care alone, SGLT2i provided an additional 0.577 QALYs and 0.809 LYs. Among available treatments, SGLT2i had the highest likelihood of being cost-effective in the Malaysian context. |
| 4 | Goh, 2023<sup>32</sup><br>Effect of empagliflozin in patients with type 2 diabetes during Ramadan on volume status, ketonaemia, and hypoglycaemia | Prospective cohort | Malaysia | Empagliflozin | Adult Muslims with T2DM<br>N: 98 participants<br>Median age: 48 (Empagliflozin), 51.5 (Control) | Aim:<br>To investigate the effects of empagliflozin use in fasting T2DM patients during Ramadan.<br>Findings:<br>The baseline measurement for body size, blood pressure and kidney function were comparable between the two groups, empagliflozin and control. No significant changes in blood pressure, body weight, urea, creatinine, eGFR or hemoglobin levels in either group. There was no significant difference observed in blood ketone levels (empagliflozin vs control, $0.17 \pm 0.247$ mmol/L vs |
|  |  |  |  |  |  | 0.13 ± 0.082 mmol/L, p = 0.304) or the incidence of hypoglycemia (empagliflozin vs control, 19.1% vs 16%, p = 0.684). |
| 5 | Wan Seman, 2016<sup>33</sup><br>Switching from sulphonylurea to a sodium-glucose cotransporter2 inhibitor in the fasting month of Ramadan is associated with a reduction in hypoglycaemia | Randomized, open-label, two-arm parallel-group study | Malaysia | Dapagliflozin | Adults with T2DM<br>N: 110 patients<br>Mean age DAPA + MET group: 53±9.1<br>SULPHONYL + MET group: 56±9.1 | **Aim:**<br>To assess the risk of hypoglycaemia associated with the use of dapagliflozin compared with that of sulphonylurea in patients with T2DM who fast during Ramadan.<br>**Findings:**<br>A small percentage of patients in the dapagliflozin group experienced hypoglycaemia compared to those in the sulphonylurea group: 6.9% (4 patients) versus 28.8% (15 patients), p = 0.002. The risk of hypoglycaemia in the fourth week of Ramadan was significantly lower in the dapagliflozin group, with a relative risk (RR) of 0.24 (95% CI: 0.09–0.68; p = 0.002). There were no significant differences between the groups in terms of postural hypotension (13.8% vs 3.8%; p = 0.210) or urinary tract infections (10.3% vs 3.8%; p = 0.277). In combination with metformin, dapagliflozin was associated with a lower risk of hypoglycaemia than sulphonylurea. |
| 6 | Goh, 2022<sup>38</sup><br>Real-world evaluation of sodium-glucose co-transporter-2 | Retrospective cohort | Singapore | Canagliflozin, Dapagliflozin, Empagliflozin | T2DM patients<br>N: 67,556 patients<br>Mean age: | **Aim:**<br>To compare the effects of SGLT2 inhibitors with DPP4 inhibitors on patient outcomes in an ethnically diverse Asian population using real-world evidence and further translate such differences into any potential healthcare cost savings. |
| | inhibitors and dipeptidyl peptidase-4 inhibitors for managing type 2 diabetes mellitus: a retrospective multi-ethnic cohort study | | | | 56.3±10.2 – 62.9±11.6 | Findings:<br>Patients who started treatment with SGLT2i were more likely to reach their glycaemic control targets compared to those on DPP4i (RR 1.09; 95% CI 1.04–1.14), a finding that was significant only among patients of Chinese ethnicity. There was no increased risk of diabetic ketoacidosis observed in those using SGLT2i. SGLT2i were also linked to a lower risk of hypoglycaemia (RR 0.69; 95% CI 0.59–0.82) and urinary tract infections (RR 0.52; 95% CI 0.43–0.63), although the reduction in hypoglycaemia was not statistically significant in Malay patients. Compared to DPP4i, SGLT2i were associated with a 12% reduction in hospitalisations for any cause and a 34% decrease in all-cause mortality, potentially leading to over \$50 million in healthcare savings over a 10-year period. |
| 7 | Low, 2022<sup>39</sup><br>Association between use of sodium-glucose co-transporter-2 (SGLT2) inhibitors and cognitive function in a longitudinal study of patients with type 2 diabetes | Prospective cohort | Singapore | SGLT2 inhibitors (Canagliflozin, dapagliflozin, empagliflozin) | Adults with T2DM<br>N: 476 patients<br>Mean age: 60.6±7.4 | Aim:<br>Explore association between SGLT2i and longitudinal changes in cognitive function in adults with T2DM and assessed the cognitive domains which were impacted by SGLT2i<br>Findings:<br>Among the 138 patients (29.0%) using SGLT2 inhibitors, 84 (17.7%) had been on the medication for less than 3 years, while 54 (11.3%) had used it for 3 years or more. In the unadjusted analysis, SGLT2i use was significantly linked to improved language scores on the RBANS cognitive test<br>(coefficient 0.60; 95% CI 0.10–1.11; p = 0.019). This positive association remained significant after adjusting for potential confounders (coefficient 0.74; 95% CI 0.12–1.36; p = 0.019). In the fully adjusted model, SGLT2i use for 3 years or longer was associated with improvements in overall RBANS scores (coefficient 0.54; 95% CI 0.13–0.95; p = 0.010) and specifically in the language domain (coefficient 1.12; 95% CI 0.27–1.97; p = 0.010). |
| 8 | Shao, 2018<sup>40</sup><br>Short-term outcomes of patients with Type 2 diabetes mellitus treated with canagliflozin compared with sitagliptin in a real-world setting | Retrospective cohort | Singapore | Canagliflozin | Adults with T2DM<br>N: 57 patients<br>Mean age: 46.4±12.1 | Aim:<br>To evaluate the effectiveness and safety of canagliflozin as compared to sitagliptin in a real-world setting<br>Findings:<br>A total of 57 patients were included in the study (22 received canagliflozin 300 mg and 35 received sitagliptin 100 mg). Baseline characteristics were comparable between the two groups, with an overall mean HbA1c of 9.4% ± 1.4%. Treatment with canagliflozin 300 mg resulted in significantly greater reductions in HbA1c (LS mean change: –1.6% vs. –0.4%; p < 0.001), body weight (–3.0 kg vs. 0.2 kg; p < 0.001), and systolic blood pressure (–9.7 mmHg vs. 0.4 mmHg; p < 0.001) compared to sitagliptin 100 mg. Around half of the patients taking canagliflozin (10 out of 22) experienced mild side effects related to osmotic diuresis, though these did not lead to treatment discontinuation. |
| 9 | Shao, 2018<sup>41</sup> | Prospective | Singapore | Canagliflozin, | Adult T2DM | Aim: |
|  | The effect of Ramadan fasting and continuing sodium-glucose co-transporter-2 (SGLT2) inhibitor use on ketonemia, blood pressure and renal function in Muslim patients with type 2 diabetes | cohort |  | Empagliflozin | patients observing Ramadan fasting<br>N: 68<br>Mean age: 52.2±11.1 | The effect of Ramadan fasting and continuing sodium-glucose co-transporter-2 (SGLT2) inhibitor use on ketonemia, blood pressure and renal function in Muslim patients with type 2 diabetes.<br>Findings:<br>A total of 68 patients with comparable baseline characteristics were enrolled in the study: 35 in the study group (on stable dose of SGLT2i) and 33 in the control group (not on SGLT2i during study period). During Ramadan fasting, both groups showed similar changes in various health parameters. There were no significant differences in weight loss (LS mean change: -1.8 kg vs. -1.1 kg; p = 0.205), estimated glomerular filtration rate (eGFR) (-6.0 vs. -4.2 ml/min/1.73 m<sup>2</sup>; p = 0.399), sitting systolic blood pressure (-8.1 vs. -10.4 mmHg; p = 0.569), sitting diastolic blood pressure (-3.7 vs. -3.5 mmHg; p = 0.934), or plasma β-hydroxybutyrate levels (-0.01 vs. -0.02 mmol/L; p = 0.649) between the groups. Ramadan fasting was associated with significant changes in weight, BP and eGFR regardless whether patients were on SGLT2 inhibitor treatment. Continued use of SGLT2 inhibitors during Ramadan did not increase ketonemia, nor increase risk of eGFR deterioration and hypoglycaemia. |
| 10 | Chanawong, 2023<sup>25</sup><br>Renoprotective Effect of Thai | Retrospective cohort | Thailand | Dapagliflozin, Empagliflozin | Patients with T2DM who have been prescribed either DPP4i or SGLT2i | Aim:<br>To evaluate the renoprotective effects of SGLT2 inhibitors compared with DPP4 inhibitors by comparing the changes in eGFR levels and the glucose-lowering effects |
|  | Patients with Type 2 Diabetes Mellitus Treated with SGLT-2 Inhibitors versus DPP-4 Inhibitors: A Real-World Observational Study |  |  |  | N: 1079 patients (388 in the SGLT2i group and 691 in the DPP4i group)<br>Median age: 64 – 66 | **Findings:**<br>A total of 388 patients were treated with SGLT2 inhibitors and 691 with DPP4 inhibitors. After 18 months of treatment, both groups showed a significant decline in estimated glomerular filtration rate (eGFR) compared to baseline. However, the reduction in eGFR was less pronounced in patients who started with a baseline eGFR below $60 \text{ mL/min/1.73 m}^2$, compared to those with higher baseline eGFR levels ($\geq 60 \text{ mL/min/1.73 m}^2$). Additionally, both groups experienced significant decreases in fasting blood glucose and haemoglobin A1c levels from baseline. Patients with $\text{eGFR} < 60 \text{ mL/min/1.73 m}^2$ who used SGLT2 inhibitors demonstrated a smaller decline in eGFR than those who used DPP4 inhibitors despite no statistical significance. |
| 11 | Uitrakul, 2022<sup>26</sup><br>The Incidence and Risk Factors of Urinary Tract Infection in Patients with Type 2 Diabetes Mellitus Using SGLT2 Inhibitors: A Real-World Observational | Retrospective cohort | Thailand | Dapagliflozin, Empagliflozin | Patients with T2DM<br>N: 853 participants<br>Mean age (95% CI): 55.7 (53.8 – 57.2) - 65.2 (64.1 - 67.3) | **Aim:**<br>To investigate the overall incidence of UTI related to SGLT2 inhibitors in Thai patients with type 2 diabetes mellitus, as well as its potential risk factors.<br>**Findings:**<br>The overall incidence of urinary tract infections (UTIs) was higher in the SGLT2 inhibitor group (33.49%) compared to the non-SGLT2 inhibitor group (11.72%). There was no significant difference in UTI rates between patients treated with dapagliflozin (34.00%) and those on empagliflozin (33.03%). Patients using SGLT2 inhibitors had a 3.7 |
|  | Study |  |  |  |  | times greater risk of developing a UTI compared to those not on these medications (95% CI: 2.60–5.29). Additionally, gender, age, and occupation were identified as significant risk factors for UTI in this study. |
| 12 | Sriphrapadang, 2020<sup>27</sup><br>Effectiveness and safety of sodium-glucose co-transporter-2 inhibitors in Thai adults with type 2 diabetes mellitus: a real-world study | Retrospective observational cohort | Thailand | Canagliflozin, Dapagliflozin, Empagliflozin | Adults with T2DM<br>N: 1159 patients<br>Mean age: 61.1±10.9 | **Aim:**<br>To assess the real-life clinical effectiveness, safety, tolerability and cardio-renal outcomes of SGLT2 inhibitors in Thai participants with T2DM.<br>**Findings:**<br>Among 1,159 participants (52.6% women; average age 61.1 ± 10.9 years; mean BMI 28.7 ± 5.2 kg/m<sup>2</sup>), 65.1% were treated with dapagliflozin, 34.3% with empagliflozin, and 0.6% with canagliflozin. The median duration of SGLT2 inhibitor use was 15 months (interquartile range: 8–23 months). Pre-existing conditions included coronary artery disease (16.5%), stroke (6.4%), heart failure (4.9%), and peripheral arterial disease (1.6%). Average HbA1c levels dropped by 0.7% (95% CI: –1.0 to –0.4) from a baseline of 8.3 ± 1.5%. After 24 months, there were significant reductions in body weight (by 2.5 kg), systolic blood pressure (by 3.5 mmHg), and diastolic blood pressure (by 2.4 mmHg). The median annual decline in eGFR was –1.3 ml/min/1.73 m<sup>2</sup>. Reported side effects included pollakiuria (7.2%), genital tract infections (2.8%), urinary tract infections (2.2%), and hypoglycemia (0.9%). No cases of diabetic ketoacidosis were observed during the study period. |
| Heart failure (7 studies) |  |  |  |  |  |  |
| 13 | Tan, 2024<sup>34</sup><br>Cost-effectiveness of empagliflozin in the treatment of Malaysian patients with chronic heart failure and preserved or mildly reduced ejection fraction | Cost-effectiveness<br>Healthcare system perspective | Malaysia | Empagliflozin | HF patients with EF>40%<br>Aged ≥ 18 years | Aim:<br>To perform a cost-utility analysis (CUA) of add-on empagliflozin to SoC versus SoC alone in EF>40% patients from the perspective of Malaysian healthcare system, and to analyse the economic value when it is prescribed for HF patients irrespective of their ejection fraction by taking into consideration the findings of the early analysis for HFrEF<br>Findings:<br>In the base-case analysis, the incremental cost-effectiveness ratio (ICER) for treating heart failure (HF) patients with an ejection fraction (EF) greater than 40% was RM 40,454 per QALY gained. Given a cost-effectiveness threshold of RM 47,439 per QALY, empagliflozin was considered cost-effective in 57% of simulations. The model's results were particularly sensitive to factors such as empagliflozin's effectiveness in reducing HF-related hospitalizations, cardiovascular mortality, and its price. For the overall HF population, the ICER was lower, at RM 29,463 per QALY gained. These findings suggest that empagliflozin is likely a cost-effective treatment for HF patients with EF >40% from the perspective of the Malaysian healthcare system. |
| 14 | Tan, 2024<sup>35</sup> | Budget Impact | Malaysia | Dapagliflozin | Malaysian HF | Aim: |
|  | Budget Impact Analysis of Dapagliflozin in Treating Patients With Heart Failure With Reduced Ejection Fraction From the Perspective of Malaysian Public Healthcare System | Analysis<br>Healthcare system perspective |  |  | patients who would be diagnosed and treated in MOH facilities<br>No mean age reported | To explore the budgetary impact of adding dapagliflozin to the SoC compared to SoC alone for treatment of HFrEF from the MOH's perspective.<br>Findings:<br>The base-case analysis projected that over a five-year period, incorporating dapagliflozin into treatment for heart failure with reduced ejection fraction (HFrEF) would lead to total cost savings of RM 2.6 million (approximately USD 0.6 million or EUR 0.5 million), amounting to a 0.3% reduction in overall costs. These savings were mainly due to decreased spending on hospitalizations for heart failure (hHF). Additionally, dapagliflozin was estimated to prevent 731 hHF events and 366 cardiovascular deaths. Sensitivity and scenario analyses indicated that the results were most influenced by assumptions related to loop diuretic usage and the price of dapagliflozin. While the first four years were projected to yield cost savings or break even, a projected 2.5% annual increase in dapagliflozin use could lead to additional costs for the Ministry of Health starting in the fifth year. Overall, adding dapagliflozin to standard care has the potential to reduce heart failure hospitalizations, cardiovascular deaths, and renal complications, offering both clinical benefits for HFrEF patients and cost savings for the Malaysian healthcare system. |
| 15 | Ong, 2023<sup>36</sup> | Cost-effectiveness | Malaysia | Empagliflozin | Heart failure with reduced EF | Aim:<br>To determine the cost-effectiveness of adding |
|  | Cost-effectiveness of adding empagliflozin to the standard of care for patients with heart failure with reduced ejection fraction from the perspective of healthcare system in Malaysia | Ministry of Health perspective |  |  | Starting age of 60 years | empagliflozin to the standard of care versus SoC alone for the treatment of patients with heart failure (HF) with reduced ejection fraction (HFrEF)<br>Findings:<br>Compared to standard care (SoC) alone, adding empagliflozin to SoC for treating heart failure with reduced ejection fraction (HFrEF) was associated with higher costs (RM 25,333 vs. RM 21,675) but also provided greater health benefits (3.64 vs. 3.46 QALYs), resulting in an incremental cost-effectiveness ratio (ICER) of RM 20,400 per QALY based on the KCCQ-CSS model. A scenario analysis using NYHA classifications yielded a higher ICER of RM 36,682 per QALY. Deterministic sensitivity analysis confirmed the model's reliability and identified the cost of empagliflozin as the key factor influencing cost-effectiveness. When government medication procurement prices were applied, the ICER dropped significantly to RM 6,621 per QALY. Overall, the results suggest that empagliflozin is likely to be a cost-effective option from the perspective of the Ministry of Health. |
| 16 | Mendoza, 2021<sup>44</sup><br>Cost-utility analysis of add-on dapagliflozin in heart failure with reduced ejection fraction | Cost-utility<br>Healthcare provider's perspective | Philippines | Dapagliflozin | HF patients with New York Heart Association class II, III, and IV whose ejection fraction was $\leq 40\%$ | Aim:<br>To determine the cost-effectiveness of dapagliflozin in addition to standard therapy versus standard therapy alone among patients with HFrEF using the public healthcare provider's perspective in the Philippines.<br>Findings: |
| | in the Philippines | | | | Starting age of 55 years | The incremental cost-effectiveness ratio (ICER) for adding dapagliflozin to standard therapy in patients with heart failure with reduced ejection fraction (HFrEF) was PHP177,868 (US\$3,434) at the current price of PHP44.00 per 10 mg tablet, and PHP160,983 (US\$3,108) at a potentially negotiated price of PHP40.00. Both ICERs fall below the cost-effectiveness threshold, which corresponds to the Philippines' 2019 per capita GDP of PHP180,500 (US\$3,485), indicating cost-effectiveness. For HFrEF patients with diabetes, the ICERs were even lower: PHP132,582 (US\$2,560) and PHP120,249 (US\$2,321), respectively, using the same price assumptions. These findings suggest that dapagliflozin is likely a cost-effective treatment option in this context. |
| 17 | Varghese, 2023<sup>42</sup><br>Cost-Effectiveness of Empagliflozin on Top of Standard of Care for Heart Failure With Reduced Ejection Fraction in Singapore | Cost-effectiveness<br>Healthcare perspective | Singapore | Empagliflozin | Heart failure with reduced EF<br>No Mean age reported | Aim:<br>To estimate the cost-effectiveness of empagliflozin + SoC versus SoC in patients with HF with reduced ejection fraction<br>Findings:<br>The combination of empagliflozin with standard care (SoC) was found to be highly cost-effective compared to SoC alone, with an incremental cost-effectiveness ratio (ICER) of less than SGD\$ 8,000 per quality-adjusted life year (QALY) gained. The base-case findings were supported by consistent results across various scenario and sensitivity analyses, confirming the robustness of the model. When health states were defined using Kansas City |
| | | | | | | Cardiomyopathy Questionnaire – Clinical Summary Score (KCCQ-CSS) quartiles, the ICER dropped further to SGD\$ 4,625 per QALY. These outcomes suggest that empagliflozin is likely a cost-effective option for treating patients with heart failure with reduced ejection fraction (HFrEF). |
| 18 | Krittayaphong 2022<sup>28</sup><br>Cost-Utility Analysis of Combination Empagliflozin and Standard Treatment Versus Standard Treatment Alone in Thai Heart Failure Patients with Reduced or Preserved Ejection Fraction | Cost-utility analysis<br>Healthcare system perspective | Thailand | Empagliflozin | Heart failure with reduced or preserved EF<br>Starting age of 60 years | Aim:<br>Cost-utility analysis of combination empagliflozin and standard treatment (ST) versus ST alone in Thai HF patients with HFrEF or HFpEF<br>Findings:<br>In patients with heart failure with reduced ejection fraction (HFrEF), adding empagliflozin resulted in a gain of 0.26 life years and 0.20 quality-adjusted life years (QALYs) at an additional cost of 409.82 USD compared to standard treatment alone, with an ICER of 69,218 THB per QALY (approximately 2,065 USD per QALY gained). For patients with heart failure with preserved ejection fraction (HFpEF), empagliflozin added 0.07 life years and 0.05 QALYs at an extra cost of 622.49 USD, resulting in an ICER of 395,826 THB per QALY (about 11,809 USD per QALY gained). These findings suggest that empagliflozin is likely a cost-effective add-on therapy for HFrEF patients but not for those with HFpEF. |
| 19 | Krittayaphong 2021<sup>29</sup> | Cost-utility analysis | Thailand | Dapagliflozin | HFrEF patients with left ventricular | Aim:<br>To determine the cost-utility of add-on dapagliflozin treatment for HFrEF |
| | Cost-utility analysis of add-on dapagliflozin treatment in heart failure with reduced ejection fraction | Healthcare system perspective | | | ejection fraction (LVEF) $\leq 40\%$, and New York Heart Association (NYHA) class II–IV with an average age of 65 years.<br>Starting age of 65 years | Findings:<br>The addition of dapagliflozin to standard therapy increased costs from 17,442 THB (559 USD) to 54,405 THB (1,745 USD), while improving quality-adjusted life years (QALYs) from 6.33 to 6.92, resulting in an incremental cost-effectiveness ratio (ICER) of 62,090 THB/QALY (1,991 USD/QALY). Sensitivity analyses showed that adding dapagliflozin was cost-effective in 87% of cases at a willingness-to-pay threshold of 160,000 THB/QALY (5,131 USD/QALY). The ICER was higher for patients without diabetes compared to those with diabetes (68,304 vs. 47,613 THB/QALY or 2,191 vs. 1,527 USD/QALY). Overall, dapagliflozin is likely to be a cost-effective treatment option for patients with heart failure with reduced ejection fraction (HFrEF). |
| Chronic kidney disease (4 studies) |  |  |  |  |  |  |
| 20 | Lim, 2024<sup>37</sup><br>Cost-effectiveness analysis of dapagliflozin for people with chronic kidney disease in Malaysia | Cost-effectiveness<br>Healthcare perspective | Malaysia | Dapagliflozin | CKD<br>No mean age reported | Aim:<br>Assessed the cost effectiveness of the introduction of dapagliflozin in addition to SoC versus SoC<br>Findings:<br>Dapagliflozin combined with standard care (SoC) was found to be the dominant treatment compared to SoC alone, with costs of RM 81,814 versus RM 85,464 (USD 19,762 vs. USD 20,644). Adding dapagliflozin for patients with chronic kidney disease (CKD) increased life expectancy by 0.46 years and quality-adjusted life years (QALYs) by 0.41 compared to SoC alone (10.01 vs. 9.55 years) |
|  |  |  |  |  |  | and 8.76 vs. 8.35 QALYs). This corresponds to a saving of RM 8,894 (USD 2,148) per QALY gained. The benefits stemmed from delaying CKD progression, which reduced the costs associated with dialysis and kidney transplantation. These findings remained consistent despite variations in disease management costs and patient subgroups and fell below the willingness-to-pay threshold of RM 46,000 per QALY. Overall, dapagliflozin is likely to be cost-saving, though confirmation through real-world data is recommended. |
| 21 | Vareesangthip 2022<sup>30</sup><br>Cost–Utility Analysis of Dapagliflozin as an Add-on to Standard of Care for Patients with Chronic Kidney Disease in Thailand | Cost-utility analysis<br>Societal perspective (direct medical and non-medical costs were included) | Thailand | Dapagliflozin | CKD<br>Starting age of 60 years | Aim:<br>To evaluate the cost–utility of dapagliflozin in addition to SoC compared with SoC alone<br>Findings:<br>Adding dapagliflozin to standard care (SoC) was estimated to increase life expectancy by 0.34 years and quality-adjusted life years (QALYs) by 0.30 compared to SoC alone (7.13 vs. 6.78 years and 5.10 vs. 4.80 QALYs). The total cost was lower with dapagliflozin treatment than with SoC alone (648,413 THB vs. 689,284 THB or 20,947.64 USD vs. 22,268.01 USD). The cost savings were primarily due to reduced expenses related to dialysis and kidney transplantation. Overall, dapagliflozin is likely to be cost-saving by delaying the progression of chronic kidney disease to dialysis. |
| 22 | Varghese, 2024<sup>45</sup> | Cost-effectiveness | Malaysia, Thailand, | Empagliflozin | CKD | Aim:<br>To assess the economic value of empagliflozin in |
|  | Cost-effectiveness of add-on empagliflozin versus standard of care in management of CKD in Malaysia, Thailand and Vietnam – findings from a modelling study assessing an EMPA-KIDNEY eligible population, using CKD progression model | (modelling)<br>Multiple countries (3x)<br>Healthcare perspective | Vietnam |  | No mean age reported | patients with CKD in Malaysia, Thailand and Vietnam<br>Findings:<br>Adding empagliflozin to standard care (SoC) was found to be cost-saving in Malaysia and Thailand, and cost-effective in Vietnam, with an ICER of 77,838,407 Vietnam Dong per QALY. This is below the willingness-to-pay threshold of 96,890,026 Vietnam Dong per QALY. Most of the cost savings over a patient's lifetime came from preventing or delaying the need for dialysis or kidney transplantation, with these savings nearly double the additional treatment expenses. The findings were consistent regardless of diabetes status and across a wide range of albuminuria levels. Overall, empagliflozin is expected to be cost-saving in Malaysia and Thailand and cost-effective in Vietnam. |
| 23 | Liu, 2022<sup>43</sup><br>A real-world study on SGLT2 inhibitors and diabetic kidney disease progression | Retrospective cohort | Singapore | Canagliflozin, Dapagliflozin, Empagliflozin | Patients with diabetic kidney disease<br>N: 4,446 patients<br>Mean age: 60.6±13.5 | Aim:<br>To evaluate the renal outcomes of CKD patients with or without SGLT2 inhibitor use in a multi-ethnic Asian population.<br>Findings:<br>This was an analysis of 4,446 participants, of whom 1,598 were treated with SGLT2 inhibitors. The use of SGLT2is was associated with a significant reduction in the progression of chronic kidney disease (CKD), with a hazard ratio (HR) of 0.60 (95% confidence interval [CI] 0.49–0.74). For<br>patients with an eGFR of $\geq 45$ mL/min/1.73 m<sup>2</sup>, the HR was 0.60 (95% CI 0.47–0.76), and for those with eGFR between 15–44 mL/min/1.73 m<sup>2</sup>, the HR was 0.43 (95% CI 0.23–0.66). Additionally, the risk of developing end-stage kidney disease (ESKD) was reduced in the entire cohort [HR 0.33 (95% CI 0.17–0.65)] and particularly in patients with eGFR 15–44 mL/min/1.73 m<sup>2</sup> [HR 0.24 (95% CI 0.09–0.66)]. Among the SGLT2 inhibitors, empagliflozin demonstrated a consistent reduction in renal risk across CKD stages 1 through 4, compared to canagliflozin and dapagliflozin. |

ASCVD: Atherosclerotic Cardiovascular Disease; BP: Blood pressure; CKD: Chronic Kidney Disease; CI: Confidence Interval; CrCl: Creatinine Clearance; CV: Cardiovascular; DAPA: Dapagliflozin; DPP4i: Dipeptidyl Peptidase-4 inhibitor; EF: Ejection Fraction; eGFR: estimated Glomerular Filtration Rate; ESKD: End-Stage Kidney Disease; GLP-1RA: Glucagon-like peptide-1 receptor agonist; HbA1c: Hemoglobin A1c; HF: Heart Failure; HFpEF: Heart Failure with Preserved Ejection Fraction; HFrEF: Heart Failure with Reduced Ejection Fraction; hHF: hospitalization for Heart Failure; HR: Hazard ratio; ICER: Incremental Cost-Effectiveness Ratio; KCCQ-CSS: Kansas City Cardiomyopathy Questionnaire Clinical Summary Score; KT: Kidney Transplantation; LS: Least Squares; LVEF: Left Ventricular Ejection Fraction; LY: Life-years; MDPI: Multidisciplinary Digital Publishing Institute; MET: Metformin; MoH: Ministry of Health; NYHA: New York Heart Association; PHP: Philippine Peso; PLOS: Public Library of Science; QALY: Quality-Adjusted Life Year; RBANS: Repeatable Battery for the Assessment of Neuropsychological Status; RM: Ringgit Malaysia; RR: Relative risk; SoC: Standard of Care; SGD\$: Singapore Dollars; SGLT2i: Sodium Glucose Co-Transporter 2 inhibitor; ST: Standard Treatment; SULPHONYL: Sulphonylurea; THB: Thai Baht; T2DM: Type 2 Diabetes Mellitus; USD: United States Dollars; UTI: Urinary Tract Infections

### Economic evaluations

Among the 13 studies that focused on the economic evaluation of SGLT2 inhibitors, 6 studies^31,34,36,37,42,45^ were cost-effectiveness analyses, 5 studies^24,28–30,44^ were cost-utility analyses, 1 study^23^ was a combination of cost-utility and budget impact analysis and 1 study^35^ was a budget impact analysis. Overall, 3 studies^23,24,30^ adopted a societal perspective as part of their evaluation, while the remaining 10 studies^28,29,31,34–37,42,44,45^ utilizing the perspectives of the healthcare systems or providers. Of these 13 studies, 12 studies ^23,24,28–31,34,36,37,42,44,45^ adopted a discount rate of 3% (in accordance with current practices ^46^ for economic evaluation). The remaining study was a budget impact analysis^35^ that was conducted without discounting as per guidelines.^47^ Discount rates are sometimes applied in economic evaluation as costs and benefits of health interventions may occur at different time points and valued differently.^48^ Economic evaluations usually consider a discount rate of 3-6%.^49^

SGLT2 inhibitors were demonstrated to be cost-effective or cost-saving in 11 out of the 13 studies,^28–31,34–37,42,44,45^ while 2 studies ^23,24^ conducted in Thailand suggested that it would not be cost-effective in the Thai context. Deerochanawong et al.^24^ conducted an economic evaluation of add-on dapagliflozin in patients with type 2 diabetes and high cardiovascular risk, and found that this was not cost-effective. Similarly, Kongmalai et al.^23^ evaluated the addition of SGLT2 inhibitors to standard of care for treating patients with type 2 diabetes and HF and found it to be not cost-effective. Kongmalai et al. proposed a price reduction for SGLT2 inhibitors by 55.6% to make these medications cost-effective.^23^ The potential reason for the inconsistent findings of these 2 studies compared to the other 11 studies could be due to the methodology. The societal perspective was applied in these 2 studies, while the healthcare system perspective was adopted in other studies. However, the third study (also conducted in Thailand) that considered a societal perspective showed that the addition of dapagliflozin was cost saving.^30^ Vareesangthip conducted a cost-utility analysis of dapagliflozin as an add-on to current therapy in patients with or without T2DM who had an eGFR of 25-75mL/min per 1.73m^2^ of body surface area and a urinary albumin-to-creatinine ratio of 200-5000mg/g.^30^ In this instance, the benefit of dapagliflozin was slowing the progression of CKD and decreasing the possibility of the need for dialysis and kidney transplantation which offset the costs associated with dapagliflozin therapy.^30^

### Safety and efficacy of SGLT2 inhibitors

Of the 23 included studies, 10 studies^25–27,32,33,38–41,43^ (9 observational studies and 1 randomized controlled trial) aimed to examine the safety and efficacy of SGLT2 inhibitors in the Southeast Asian populations.

The common side effects of SGLT2 inhibitors reported in these studies were genitourinary infections, like urinary tract infections, hypoglycemia and potential dehydration. Uitrakul and colleagues conducted a study in 853 patients with type 2 diabetes in Thailand and found that the incidence of urinary tract infection was significantly higher in those who used SGLT2 inhibitors than non-users (33.5% vs 11.7%).^26^ Regarding dehydration and hypoglycemia, most studies concluded that the risks of these side effects were not increased in SGLT2 inhibitor users compared to non-users, even during fasting periods.^32,33^ SGLT2 inhibitor use was also not associated with increased risk of diabetic ketoacidosis during Ramadan fasting.^32,41^

The only randomized controlled trial in this review aimed to assess whether switching from sulfonylurea to a dapagliflozin in the fasting month of Ramadan resulted in a reduction in hypoglycemia.^33^ The study reported that fewer patients exhibited hypoglycemia in the dapagliflozin group than in the sulfonylurea group.^33^

Regarding the clinical benefits, a study in Singapore reported an association between ≥ 3 years of SGLT2 inhibitor use and improved cognitive scores.^39^ Two studies in Singapore and Thailand reported that the use of SGLT2 inhibitors was more effective than non-use in reducing HbA1c, body weight and systolic blood pressure in patients with type 2 diabetes.^27,40^ Similar findings were reported in another study in Singapore where SGLT2 inhibitor initiation was associated with significantly (p<0.001) lower mean HbA1c than those initiated on DPP4- inhibitors, although the difference (7.54% vs 7.68%) was modest.^38^

SGLT2 inhibitor initiation was also associated with fewer hospitalizations and deaths up to one-year post-initiation.^38^ A study of 4,446 patients with type 2 diabetes in Singapore ^43^ found that SGLT2 inhibitors demonstrated protective effects on chronic kidney disease progression, with the hazard ratio (HR) and 95% confidence interval (CI) for developing end stage kidney disease of 0.33 (0.17 - 0.65), compared to non-use of SGLT2 inhibitors.

## Discussion

We have conducted the first systematic review of studies examining the use of SLGT2 inhibitors among adults living in Southeast Asian countries. Our findings indicate that evidence related to the use of SGLT2 inhibitors is still limited in this region. Previous studies have focused on the cost-effectiveness, efficacy and safety of SLGT2 inhibitors. There were no studies that examined the availability of these medications in the region. Limited evidence, from just 5 out of 11 countries in the region (Malaysia, Philippines, Singapore, Thailand and Vietnam), suggests that SGLT2 inhibitors appear to be safe, effective, and are likely to be cost-effective but inconsistent findings exist across populations. The lack of robust evidence underscores a significant gap in our understanding of access to SGLT2 inhibitors in the region.

Although evidence suggests that the cardiovascular and renal benefits of SGLT2 inhibitors observed in global trials may be generalizable to Southeast Asian populations^50^, further local studies are necessary to confirm these findings. There has been evidence of the influence of racial, ethnic and regional groups on the effect of SGTL2 inhibitors. A 2024 meta-analysis of 7 trials comparing SGLT2 inhibitors vs placebo reported consistent benefits observed among White and Asian populations, and a lack of benefits in Black and other populations: HRs for major adverse cardiovascular events were 0.92 (95% CI 0.86 – 0.98) among White participants, 0.69 (95% CI 0.53 – 0.92) among Asian participants, 1.11 (95% CI 0.82 – 1.51) among Black participants, and 0.83 (95% CI 0.52 – 1.35) among other race.^51^ A 2025 meta-analysis of 14 randomized controlled trials of 94,445 participants demonstrated that SGLT2 inhibitors showed consistent efficacy on the studied outcomes (composite of cardiovascular death or heart failure hospitalization, composite of cardiovascular disease and chronic kidney disease progression, major adverse cardiovascular events, cardiovascular death, all-cause death) across three pre-specified racial groups of Asian, Black and White, except for heart failure hospitalization. ^52^ The efficacy of SGLT2 inhibitors on heart failure hospitalization was more pronounced in Black and Asian patients compared to White patients.^52^. However, there were no studies from Southeast Asian countries in these meta-analyses.

This review revealed that there have been limited studies on SGLT2 inhibitors in older adults in Southeast Asia, and there has been no evidence on the use of this drug class in those with frailty. Further studies are needed to provide region-specific data on the safety and efficacy of SGLT2 inhibitors in these populations. Evidence from the literature so far has shown that SGLT2 inhibitors are effective and safe in frail patients compared to non-frail patients. Post-hoc analyses from major trials on SGLT2 inhibitors, such as the DAPA-HF ^53^, DELIVER ^54^, EMPEROR-Preserved (Empagliflozin Outcome Trial in Patients With Chronic Heart Failure With Preserved Ejection Fraction) ^55^, DAPA-CKD (Dapagliflozin and Prevention of Adverse Outcomes in Chronic Kidney Disease) ^56^, the CANVAS program and the CREDENCE trial, ^22^ showed that the benefit of these medications was consistent across the spectrum of frailty, with no significant difference in safety. However, most of the trial participants were recruited from more developed countries, limiting generalizability to the local settings in many low- and middle-income countries in the Southeast Asia. This uncertainty can be a challenge for local clinicians to make evidence-informed decisions accounting for the genetic, environmental and healthcare differences.

To address these gaps, targeted local clinical trials and more observational studies are needed to further evaluate both the therapeutic benefits and safety profiles of SGLT2 inhibitors, especially in older populations in Southeast Asia. This can include pharmacokinetic and pharmacodynamic analyses, as variations in metabolism and body composition may influence drug response. Furthermore, regionally adapted clinical guidelines and prescribed education are important to support cautious and individualized use. Without this, the growing population of older adults with diabetes in Southeast Asia may be underserved, potentially missing out on meaningful reductions in cardiovascular and renal morbidity, and further exacerbating health inequities.

Preliminary findings showed that SGLT2 inhibitors seem to be cost-effective for patients in the region. Economic evaluation of new and useful medicines likes SGLT2 inhibitors are important, particularly in areas of limited resources that are sometimes faced by low- to middle-income countries. Despite the clear clinical benefits of SGLT2 inhibitors, several barriers hinder their widespread adoption in Southeast Asia. One of the most significant challenges is the cost of these medications^19^, which limits patient access. The upcoming availability of generic SGLT2 inhibitors may help reduce access barriers by providing a more affordable cost. For example, generic versions of dapagliflozin and empagliflozin were recently made available in Malaysia. Another challenge is the uneven healthcare infrastructure across Southeast Asia. Access to healthcare and medications varies greatly between urban and rural areas, with rural populations often experiencing limited access to essential healthcare services, including regular monitoring of renal function, a crucial consideration when using SGLT2 inhibitors. Inadequate healthcare infrastructure, coupled with a lack of healthcare worker education regarding the newer pharmacological treatments^20^, may contribute to underutilization or suboptimal use of SGLT2 inhibitors in clinical practice.

In economic evaluations in healthcare, analysis is conducted from a viewpoint or perspective. Examples of these perspectives can include patient, healthcare provider, health system or societal. There are variations in methodological approaches and applications of the different perspectives. The societal approach is the broadest perspective and includes all healthcare related costs and potentially other indirect costs as well.^57^ There is no one-size-fits-all as to which perspective should be used as it has its merits, depending on the intended use of the analysis and who is it for (eg. policymaker, context).^57^ For Thailand, the Thai Health Technology Assessment guidelines recommend using a societal perspective for economic evaluations.^58^ With the increasing prevalence of diabetes and its impact in Thailand and the broader Southeast Asian region^13^, policymakers can further explore innovative ways for financing to further help improve access to SGLT2 inhibitors.

Despite existing challenges, there are substantial opportunities to increase the use of SGLT2 inhibitors in Southeast Asia. As economic development continues in many countries in the region, access to healthcare and modern medications is expected to improve. Increased investment in healthcare infrastructure, particularly in rural and underserved areas, could facilitate better management of diabetes and related comorbidities, improving access to SGLT2 inhibitors. Furthermore, as the prevalence of T2DM, chronic kidney disease, and cardiovascular disease continues to rise in Southeast Asia^59^ there is an urgent need for effective interventions that target both glycemic control and cardiovascular risk reduction. Additionally, the potential usefulness of SGLT inhibitors for improvement of non-alcoholic fatty liver disease provides another opportunity given that the region is experiencing an increasing trend of obesity.^60^ Countries in the Southeast Asian region have been reported to have some of the fastest growing rates of obesity and metabolic syndrome globally^61^ as such, the increased use of SGLT2 inhibitors offers much possibilities.

SGLT2 inhibitors, with their demonstrated efficacy in reducing cardiovascular morbidity and mortality, may become an integral part of the therapeutic arm in managing T2DM and heart failure in this region. Enhanced awareness campaigns for healthcare providers and patients about the benefits of SGLT2 inhibitors could promote their use. Finally, future clinical trials specifically designed to assess the effectiveness and safety of SGLT2 inhibitors with a substantial number of participants in Southeast Asian countries are essential. These studies would provide more granular data on ethnic-specific responses to SGLT2 inhibitors, helping to refine treatment guidelines and inform clinical practice in the region.

### Strengths and limitations

To the best of our knowledge, this is the first systematic review to summarize the evidence of research on SGLT2 inhibitors among adults living in Southeast Asian countries. The literature search conducted across three large databases, ensured a comprehensive collection of studies. The inclusion criteria and methodologies applied were designed to minimize bias and maximize the relevance of the studies obtained. However, our study has several limitations. The literature search was limited to studies published in English, omitting studies published in local languages. There may be inconsistencies among the reviewers during the study selection and data extraction phases. To address this, we have adhered strictly to the review protocol and maintained regular communications and cross-checks among team members to reduce any potential discrepancies.

## Conclusion

This study highlighted the need for further studies on the use of SGLT2 inhibitors in Southeast Asian population. Limited evidence suggests that SGLT2 inhibitor treatment may offer promising benefits for patients with type 2 diabetes, heart failure and chronic kidney disease in the region. Further research is needed to understand the availability and affordability, as well as the safety of SGLT2 inhibitors, taking into consideration the effect of ageing and frailty in this region. In addition, region-specific factors such as genetic variations, healthcare infrastructure, cost, and cultural considerations should be addressed to optimize their use. With ongoing research, improved healthcare delivery, and greater access to medications, SGLT2 inhibitors have the potential to significantly impact the treatment of T2DM and related comorbidities in Southeast Asia.

## Data Availability

All data produced in the present study are available upon reasonable request to the authors

